# A newly emergent N1 neuraminidase associated with clade 2.3.4.4b highly pathogenic avian influenza A(H5) viruses in North America

**DOI:** 10.64898/2026.03.09.26347929

**Authors:** Matthew J. Wersebe, Nicole M. Paterson, Norman Hassell, Xiao-yu Zheng, Benjamin Rambo-Martin, Julia C. Frederick, Kristine A. Lacek, Amanda H. Sullivan, Marie K. Kirby, Rebecca Kondor, Yunho Jang, Sabrina Schatzman, Han Di, C. Todd Davis

**Affiliations:** Influenza Division, National Center for Immunizations and Respiratory Diseases, US Centers for Disease Control and Prevention, Atlanta, GA USA

**Keywords:** Highly Pathogenic Avian Influenza, Neuraminidase, Phylodynamics, Protein Structure

## Abstract

We investigated the evolutionary history of the newly emergent neuraminidase (am4N1) associated with the D1.1 and D1.2 genotypes of highly pathogenic avian influenza A(H5N1) viruses in North America. Phylogenetic inference places am4N1 in a sister clade to Eurasian avian, swine, and human A(H1N1)pdm09 viruses and distinct from 1918, pre-2009 human seasonal, and classical swine A(H1N1) lineages. Am4N1 descends from diverse avian N1 genes endemic to the Americas. Phylodynamic analysis indicates a monophyletic am4N1 lineage with numerous introductions of viruses carrying the am4N1 gene likely originating from western Canada into the United States during emergence of the D1.1 and D1.2 genotypes. The lineage has diversified and accumulated deletions in the stalk domain. Despite amino acid divergence, structural modeling shows conserved neuraminidase architecture in the globular head. Given its distinct ancestry and amino acid sequence, further studies are needed to assess cross-reactivity of antibodies from prior human A(H1N1)pdm09 infections.

## Introduction

Highly pathogenic avian influenza (HPAI) viruses of A(H5) clade 2.3.4.4b have circulated widely in wild birds since their emergence in China in 2016 [1]. Since 2021, Eurasian-origin clade 2.3.4.4b A(H5N1) viruses have circulated in the Americas when they were introduced to North America likely from migratory birds [2]. This has resulted in a widespread epizootic of A(H5N1) and other A(H5Nx) viruses with unprecedented mortality in wild birds and the depopulation of more than 100 million commercial poultry in the United States in 2024 alone [3]. There have been numerous spillover events from birds to wild mammalian species throughout North America, often leading to systemic infection and death in wild mammalian hosts [4, 5].

Since their emergence in North America, clade 2.3.4.4b A(H5N1) viruses have diversified via reassortment with other low pathogenic avian influenza (LPAI) viruses [6]. Beginning in late 2024, wild birds infected with two novel genotypes of A(H5N1) were discovered along the Pacific Coast of the United States and Canada and were designated as D1.1 and D1.2 by the U.S. Department of Agriculture (USDA) [7]. These viruses were notable in that they contained a new neuraminidase (NA) gene that was different from other contemporary HPAI A(H5N1) viruses previously isolated from wild birds in North America [8]. Concomitant with the discovery of D1.1 and D1.2 genotype A(H5N1) viruses in wild birds, these viruses spilled over into commercial poultry in the Pacific Northwest region of the U.S. and western Canada. Since the initial spillover, D1.1 genotype viruses caused sporadic human infections in the U.S. in people with close contact with infected livestock and poultry on farms and in backyard settings [9]. Beginning in October 2024, the first human cases of genotype D1.1 infection were reported in Washington, U.S. poultry workers [10]. Among these poultry workers in Washington, all individuals recovered. In November 2024, public health authorities in British Columbia, Canada reported a case of D1.1 A(H5N1) in a teenager with no known exposure to infected birds or other animals. The patient experienced severe disease [11]. In December 2024, Louisiana, United States public health officials identified a D1.1 infection in an adult individual who had close contact with a backyard poultry flock infected with D1.1, the first reported human infection in the U.S. derived from a backyard flock [9]. The Louisiana patient experienced severe disease and died, the first human fatality associated A(H5N1) infection in the United States. Additionally, D1.1 caused severe disease requiring hospitalization in a second individual in Wyoming, United States in February 2025 after exposure to an infected backyard flock. In April 2025, a child in Durango, Mexico also passed away after infection with a D1.1 A(H5N1) virus [12]. Since late 2024, D1.1 has spread widely in the United States, Canada, Mexico and as far south as Guatemala being observed in all North American flyways. Further complicating matters, D1.1 A(H5N1) has caused distinct spillover events in US dairy cattle in Nevada, Arizona, and Wisconsin in January, February and December 2025, respectively, leading to an infection in a Nevada dairy worker [9]. Other recent clade 2.3.4.4b A(H5N1) human infections in North America, especially those associated with genotype B3.13, which was identified in dairy cattle in early 2024, have been largely mild. B3.13 genotype viruses have an NA gene derived from Eurasian avian N1 lineage [7].

The D1.1 and D1.2 genotypes of A(H5N1) are notable in that these viruses carry a unique N1 neuraminidase (NA), which was reported to be most like North American N1s previously isolated from wild birds and have been designated as am4N1 by the USDA. Here we investigated the phylogenetic relationships of the am4N1 lineage in the context of the global diversity of N1 NA sequences. We provide important context about the recent evolutionary history of the am4N1 NA lineage, including the hemagglutinin type diversity previously observed with similar NA proteins, phylogeographic characteristics and stalk domain changes observed. Additionally, we predict the structure of am4N1 NAs and compare these with other diverse N1 NAs and contrast the amino acid sequence identity in the primary NA protein domains. This work provides evolutionary and functional context for the ongoing epizootic and evolving pandemic threat posed by A(H5N1) viruses globally and in North America, specifically.

## Materials and Methods

Extensive details on the acquisition, isolation, and initial sequence characterization of human samples received by CDC have been previously reported in Rolfes et al. [9].

### Sequence Searches

Using the NA sequence from A/Washington/255/2024 (an early D1.1 isolate) as a reference, we conducted a search of the Global Initiative to Share All Influenza Data (GISAID) EpiFlu sequence databases for all NA N1 sequences and calculated a pairwise Tamara & Nei [13] (TN93) genetic distance. We retained all sequences with a genetic distance less than 0.1 compared to A/Washington/255/2024. Next, we supplemented am4N1-like sequences available from GISAID by downloading and assembling available D1.1 and D1.2 next-generation sequencing data from NCBI’s Sequence Read Archive (SRA) relating to ongoing HPAI influenza surveillance in the US using the IRMA assembler [14], (hereafter referred to as am4N1-like). Finally, we downloaded all NA-N1 sequences from the GISAID EpiFlu database. We limited our query to include only data with a complete CDS for NA (1410 nt), less than 5 IUPAC ambiguities and no unresolved (N) sites. We created a non-redundant sequence database of NA-N1s at the 98% sequence identity level using CD-Hit (16) to limit the number of sequences for phylogenetic inference (hereafter referred to as Global N1). Additionally, we recorded the genoflu [6, 7] segment lineage for each retained NA sequence for both datasets.

### Maximum Likelihood Phylogenetics

We extracted aligned NA sequences from DIAS-ribosome [15] and we inferred phylogenetic trees for both the am4N1-like and the Global N1 datasets using IQtree2 (v2.3.6) [16]. We determined the DNA substitution model of best fit using the Bayesian information Criteria (BIC) in IQtree2 using ModelFinder [17] for the global dataset and inferred an initial ML tree using GTR for the am4N1-like dataset. Finally, we estimated branch support for the inferred topologies using 1000 Ultrafast bootstraps [18]. For the am4N1-like dataset, we used the maximum-likelihood phylodynamic tool kit available in treetime (v0.11.4) [19] to reconstruct the evolutionary history of the am4N1 lineage as implemented in augur (v29.0.0) [20]. Within the am4N1 clade we searched for sequences in our alignment showing deletions in the stalk domain. Finally, we rooted the Global N1 phylogenetic tree to the approximate mid-point using augur refine [20].

### Protein Secondary Structure Prediction, Alignment and Distance Calculations

A subset of global N1-NA diversity was selected from GISAID to investigate structural differences apparent between the various NA lineages (Table S1). We aligned amino acid translations with MUSCLE using 8 iterations and clustering by similarity and calculated an amino acid distance matrix within the Geneious prime platform [21]. We then submitted the NA sequences to the AlphaFold3 server for protein structure prediction [22]. AlphaFold3 utilizes deep learning techniques to predict protein structures based on amino acid sequences, providing high-confidence structural models. Following structural prediction with AlphaFold3, we extracted and relaxed the head domain containing the region with the highest confidence based on model prediction, pLDDT > 90 (sites 80-479) using the Rosetta (v3.14) relax protocol as described and detailed in Navion et al. [23] and Conway et al. [24]. The relax protocol implements an energy minimization technique that refines the predicted structures by alleviating steric clashes and optimizes hydrogen bonding networks, thereby enhancing the accuracy of analysis done on structural predictions. Next, we performed pairwise alignment between the relaxed structures and recorded biophysical features using Rosetta following methodologies outlined and detailed in [25, 26]. We averaged the Root Mean Square Deviation (RMSD) values across triplicates of predicted protein structures to quantify structural differences between the relaxed models and the reference structure. The RMSD provides a measure of the average distance between atoms of superimposed proteins, serving as an indicator of conformational similarity. We compared the amino acid sequence identities within the transmembrane, stalk, and head domains of sequences that underwent structural predictions to understand how amino acid differences are partitioned throughout the protein.

## Results

### Sequence Searches

Our searches of publicly available sequences resulted in 2061 sequences for the am4N1-like dataset with TN93 genetic distances less than 0.1. Meanwhile, our global N1 sequence search returned 136,205 sequences. Clustering at 98% sequence identity yielded 844 representative sequences for global NA-N1 phylogenetic inference.

### Global NA-N1 Phylogenetics

Automated execution of Modelfinder determined that the NA sequence data fit the GTR+F+R7 substitution model best according to BIC. It is important to note that the lineage descriptors used below (e.g., A.1, B.2, etc.) are not meant to represent a comprehensive taxonomy of N1 neuraminidases but are instead a practical shorthand to refer to specific clades and lineage emergent within the phylogeny inferred from this data set. The mid-point rooted phylogeny of subsampled NA-N1 diversity showed two emergent groups within global N1 which we denote as groups A and B (Figure 1A). Group A consists of two lineages; lineage A.1, encompassing 1918 A(H1N1) and pre-2009 human seasonal A(H1N1) viruses, and lineage A.2 encompassing the classical A(H1N1) swine influenza NA, which has been largely restricted to North America with occasional detections in Eastern Asia and Europe (Figure 1A, Supplemental Data file 1). Group B N1s have greater phylogenetic diversity, consisting of 6 lineages including present day A(H1N1)pdm2009, Eurasian avian N1 (including other A(H5N1)), and Eurasian swine-associated viruses. B.1, B.2, and B.4 are small lineages with between 2 (B.2 and B.4) and 6 (B.1) retained sequences and are restricted to avian hosts. B.3 consists of 37 retained sequences descending from A/Chicken/Scotland/1959, which later gave rise to am4N1 (Figure 1B). Group B.5 consists of avian

**Figure 1.**
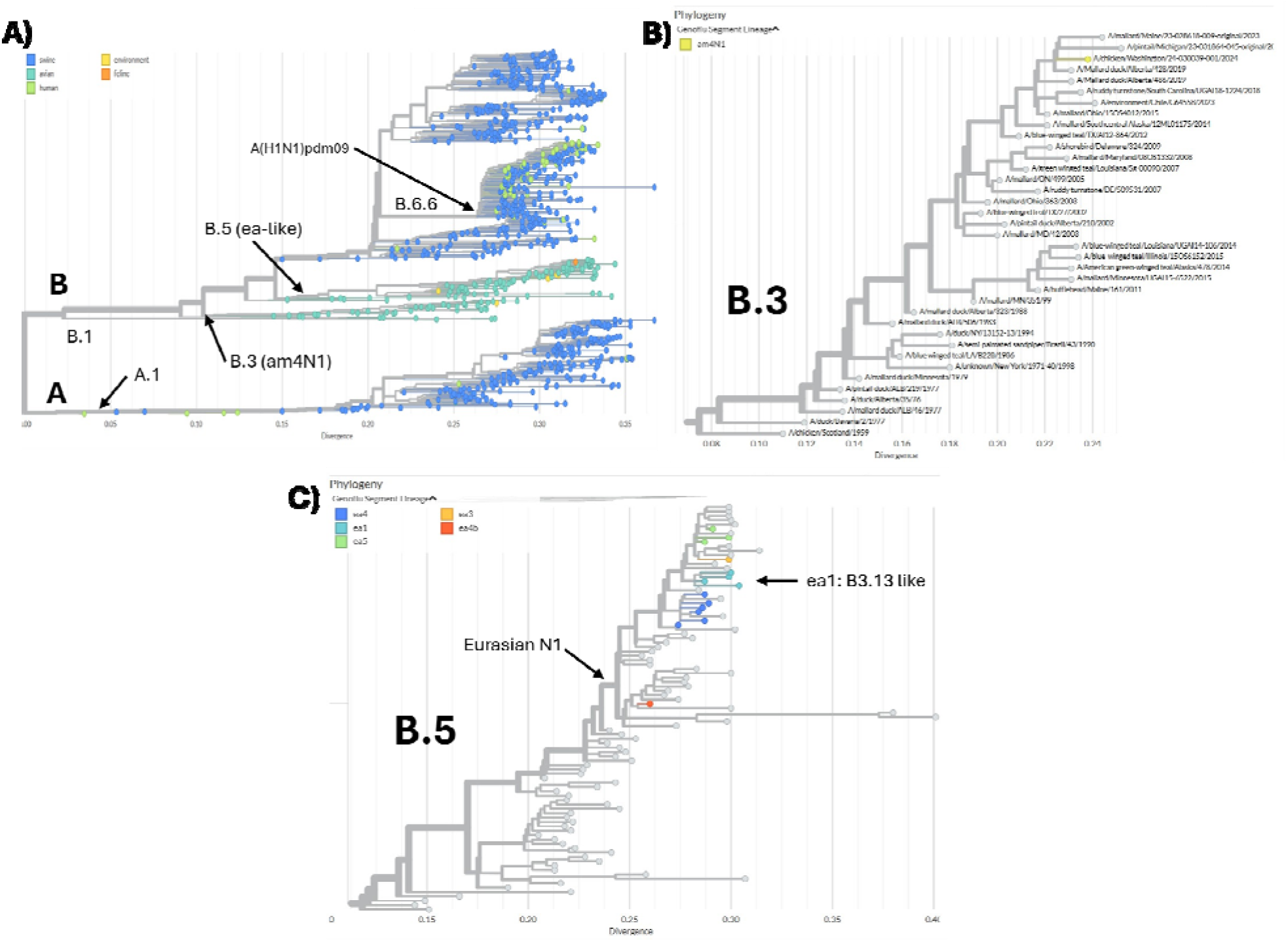
A) Mid-point rooted Phylogeny of Global N1-NA Diversity, branch lengths in units of genetic divergence. Tips are colored by generic host categories. Clade labels show the two N1 supergroups (A and B) and subclades of interest. B) Subtree showing group B.3 which includes am4N1 and its progenitors. Tips colored by *Genoflu* segment lineage assigned, grey tips are not assigned a genoflu segment lineage type. C) Subtree showing group B.5 which include Eurasian N1s associated with the initial colonization of North America by A(H5N1) HPAI. EA1 subgroup includes genotype B3.13 viruses circulating in US dairy cattle.

NA genes that include the present N1s associated with Eurasian HA clade 2.3.4.4b HPAI introductions to the Americas (Figure 1C). Group B.6 NAs are largely restricted to Eurasian swine and its derivatives. This group is highly diverse and geographically structured including seven subclades, one of which B.6.6 is the 2009 pandemic lineage. An interactive auspice build of this analysis is provided as supplementary data file 1.

### Am4N1-like Phylodynamics

NA sequences with TN93 genetic distance of less than 0.1 compared to A/Washington/255/2024 ranged in date from 1976 to the present (Table 1). Rooting of the tree estimated that all diversity within the retained sequences coalesced by July 1975 (TMRCA 1975-07-06, 1973-09-29 to 1975-12-12 95%CI;Supplement Data file 2). Root to tip regression in treetime suggested a substitution rate of 2.79 x 10^−3^ nucleotide mutations per site per year for this group, suggesting a strong temporal signal in the data (Figure S1). Contemporary am4N1-like sequences form a single monophyletic clade with 100% bootstrap support, this group diverged from other avian N1s in early 2023 (TMRCA 2023-01-25, 2022-02-13 to 2023-08-28 95%CI; Figure 2A) and can be defined by an amino acid substitution at NA:S82P along with nucleotide mutations at T244C and C588T (Figure 2B). Due to its origin in wild bird reservoirs, there were no detections of am4N1-progenitors before the outbreak began, and it was likely under-sampled from late 2019 until late 2023. In late 2024, with its reassortment with HPAI A(H5) clade 2.3.4.4b HA, the am4N1 NA was observed in western Canada but has since spread across all North American flyways with multiple introductions into the lower 48-states of the US most likely from western Canada. An interactive auspice build with data is available in supplementary data file 2.

**Table 1.** Viruses with NA sequences related to D1.X am4N1 NAs. Accession, GISAID EPI_ISL number from EpiFlu database. Location, Continent; Country; State/Province. NA, North America. A/Cackling Goose/BC/AIVPHL-2521/2024 represents the first D1.1 genome in public databases.

| Accession<br>(EPI_ISL) | Virus Name | HA | Date | Location | Phylogenetic Position | TN-93 Distance |
| --- | --- | --- | --- | --- | --- | --- |
| 8827 | A/mallard duck/ALB/26/1976 | H3 | 1976-08-11 | NA; CAN; AB | Near Root | 0.0997 |
| 294067 | A/green-winged teal/Mississippi/16OS6075/2016 | H1 | 2018-02-06 | NA; USA; MS | Ancestral am4N1 | 0.0188 |
| 19175579 | A/mallard/Connecticut/23-028159-003/2023 | H1 | 2023-06-03 | NA; USA; CT | Ancestral am4N1 | 0.011 |
| 19070538 | A/mallard/Alberta/69/2024 | H1 | 2023-04-30 | NA; CAN; AB | Pre-outbreak am4N1 | 0.009 |
| 19533299 | A/mallard/BC/AIVPHL-2360/2024 | H1 | 2024-08-20 | NA; CAN; BC | Outbreak am4N1 | 0.004 |
| 19716111 | A/mallard/Alberta/520/2024 | H1 | 2024-08-22 | NA; CAN; AB | Outbreak am4N1 | 0.006 |
| 19533311 | A/Cackling Goose/BC/AIVPHL-2521/2024 | H5 | 2024-10-29 | NA; CAN; BC | Outbreak am4N1 | 0.005 |

**Figure 2.**
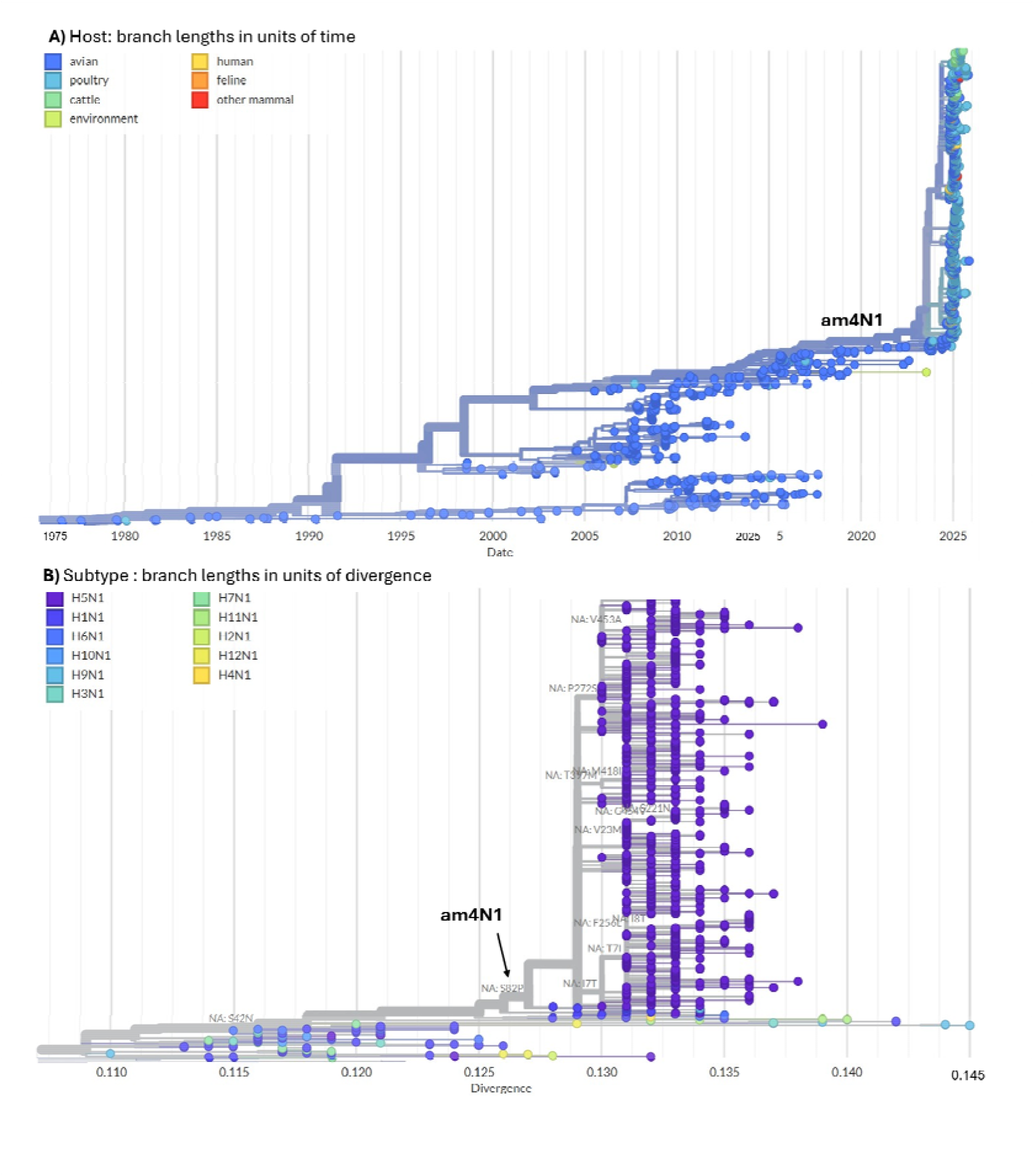
Phylodynamics of am4N1-Like NAs. Current am4N1 sequences involved in the outbreak are labelled near top of both trees. A) Time-scaled phylogenetic tree with tips colored according to the identity of virus host. B) Divergence-scaled subtree with tips colored according to the subtype associated with the virus isolate. Tree view zoomed into the am4N1 clade; branches labelled with amino acid substitutions.

### Stalk Domain Deletions in am4N1

There are seven instances of stalk deletions in the am4N1 clade (Table 2). Stalk deletions range in size from 84 bases (27 AA residues) to 51 bases (17 AA residues). The deletions are consistently observed in wild bird hosts suggesting that these are present in the wild bird population (supplemental data file 2). Only two of the 6 deletions are observed in farmed animals (table 2, rows 1 and 5). Reconstruction of the ancestral nodes leading to the terminal branches often included a high likelihood of poultry (e.g., gallinaceous host identity) on the branches ancestral to the observed deletions. One deletion, SΔAA:53-78 appears in two separate albeit closely related subclades in the tree, suggesting convergent evolution for this deletion. The stalk deletion identified in A/San Marcos/FLU-383/2025 (EPI_ISL_20254739) and A/Izabal/FLU-340/2025 (EPI_ISL_20180701) is more complex than other instances spanning a single large region along with several discontinuous codons (table 2).

**Table 2.**
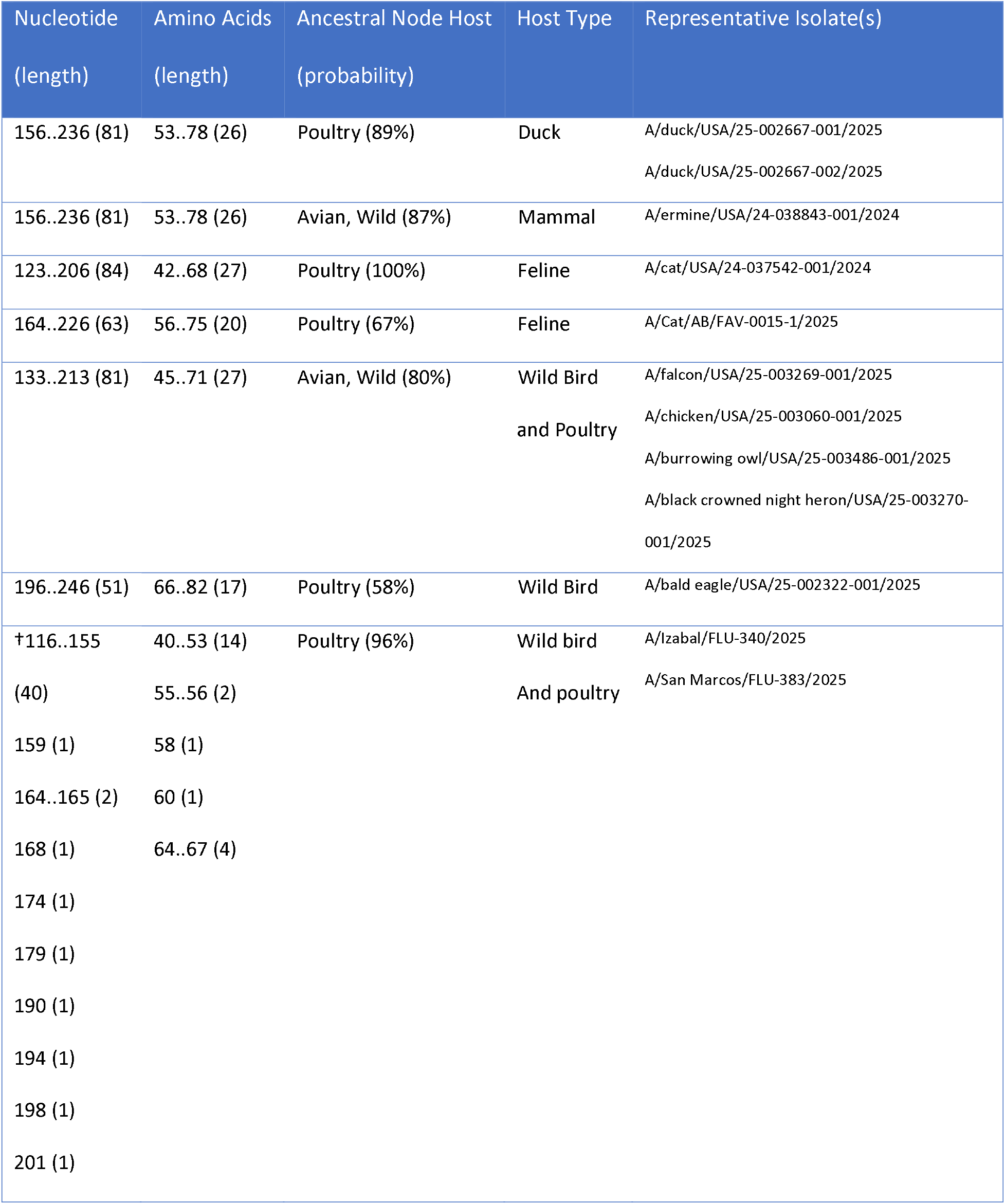
Stalk deletions in the am4N1 outbreak clade. Nucleotide positions deleted, their length in bases, amino acid positions deleted and lengths in residues, the host type of the representative isolates, and viruses with deletions. †The complexity of this deletion may be an artifact of alignment uncertainty in this region with many gaps.

### Related Viruses

Am4N1 NA is descended from NAs associated with nearly all HA types including American avian A(H1) (most common), A(H3), A(H6) and A(H10). The reassortment event leading to D1.1 and D1.2 viruses likely resulted from A(H5) co-infection in an avian host with an A(H1), as there is a recent am4N1 NA sequence, A/mallard/BC/AIVPHl-2360/2024, subtyped as an A(H1N1). Other viruses positioned as outgroups to the am4N1 clade are also A(H1N1) viruses (Table 2). A/mallard/BC/AIVPHl-2360/2024 is ancestral to all D1.1 am4N1s in North America. Thus, am4N1-NAs are likely derived from avian A(H1) circulating in North American migratory waterfowl as it is commonly detected with this HA type throughout its very recent evolutionary history (Figure 2B).

### Amino Acid Differences and Structural Predictions

NAs encode a protein that contains 4 different domains including a conserved cytoplasmic tail, a transmembrane region, the stalk domain, and a globular head which contains the enzymatic active site (Figure 3-Top Left). We retained just the globular head and stalk domains for prediction with AlphaFold, as transmembrane domains are difficult to predict with high confidence. Overall, amino acid sequence differences are greatest between A(H1N1)pdm09 NAs (CA/07/2009 & WI/67/2022) and all avian-like NA including Eurasian and am4N1s (Figure 3B; ∼50 & ∼66 AA residues). Additionally, am4N1 are divergent from the Eurasian NAs with the sequences differing at approximately 30-50 residues depending on the length of stalk deletions (Figure 3B). These differences are concentrated in the head domain (Figure 3C; positions 80-469). RSMD averaged over three replicates revealed that protein structure is largely conserved across NA-N1 diversity. For N1 sequences examined here, the greatest conservation is observed within host associations where avian sequences are most structurally like other avian NAs (Figure S3). Using the structural predictions, we calculated the relative surface accessibility of each residue in the protein and extracted the surface exposed residues that are within a 25 Å radius of the enzymatic active site. While there is a high level of conservation in this region between the N1 NA, am4N1s have a single N221S (N2 numbering N220) amino acid substitution (Figure 4) compared to N1pdm and N1-ea1. This position has been identified as a site of immune escape in previous studies but no data are currently available that assess the impact of this change on viruses expressing NA from the am4N1 lineage [27].

**Figure 3.**
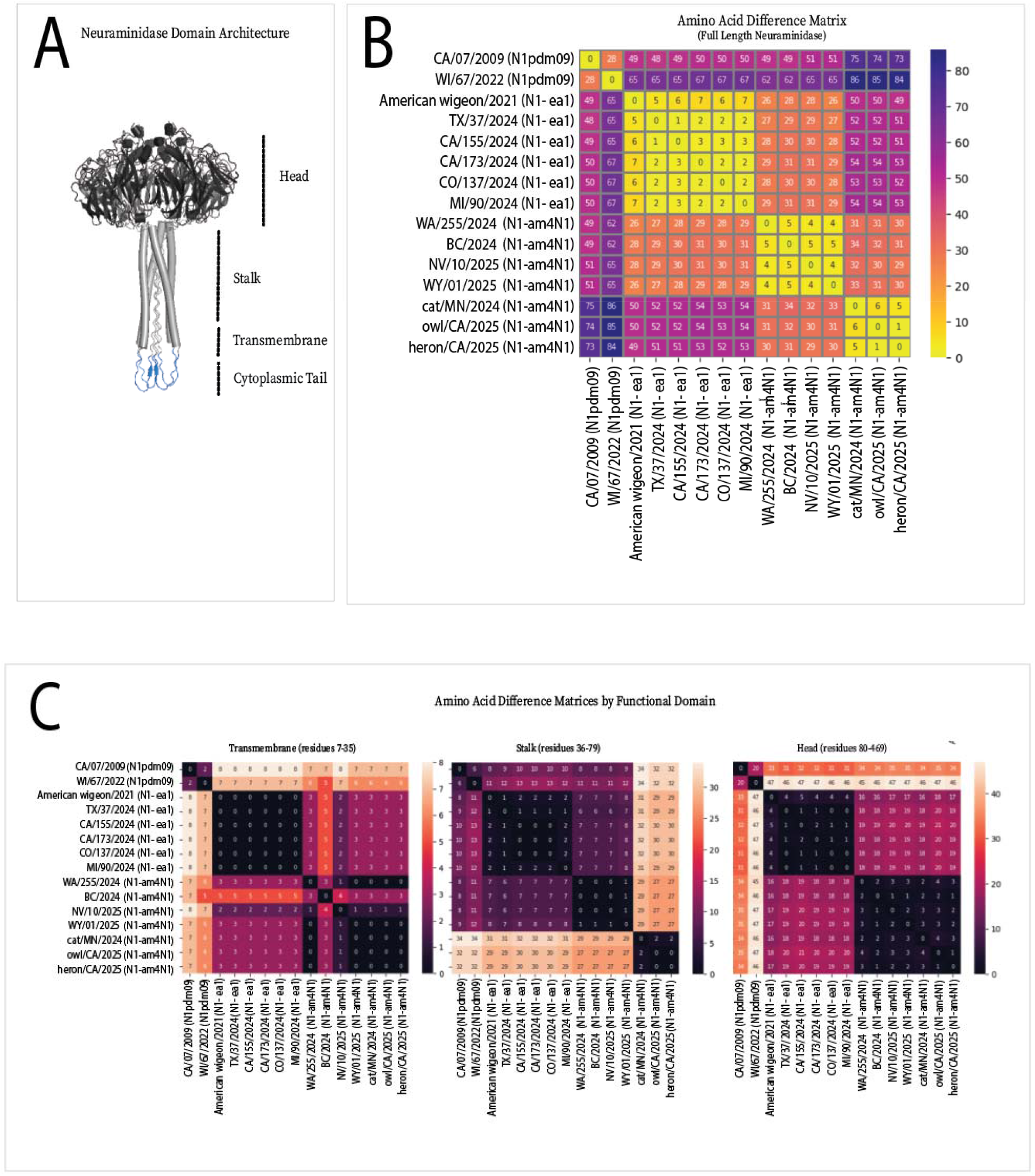
A) AlphaFold produced schematic of NA-N1 protein with domain locations (domain definitions taken from 28) noted. B) Overall amino acid distance matrix heat map across entire sequence. Numeric values are pairwise comparison of counts of amino acid differences. C) Amino acid distance matrix heat maps by protein domain excluding the highly conserved cytoplasmic tail which contained no differences across isolates. A(H1N1)pdm09 reference sequences were selected due to presence in previous seasonal influenza vaccine formulations. See Supplement for full sequence information.

**Figure 4.**
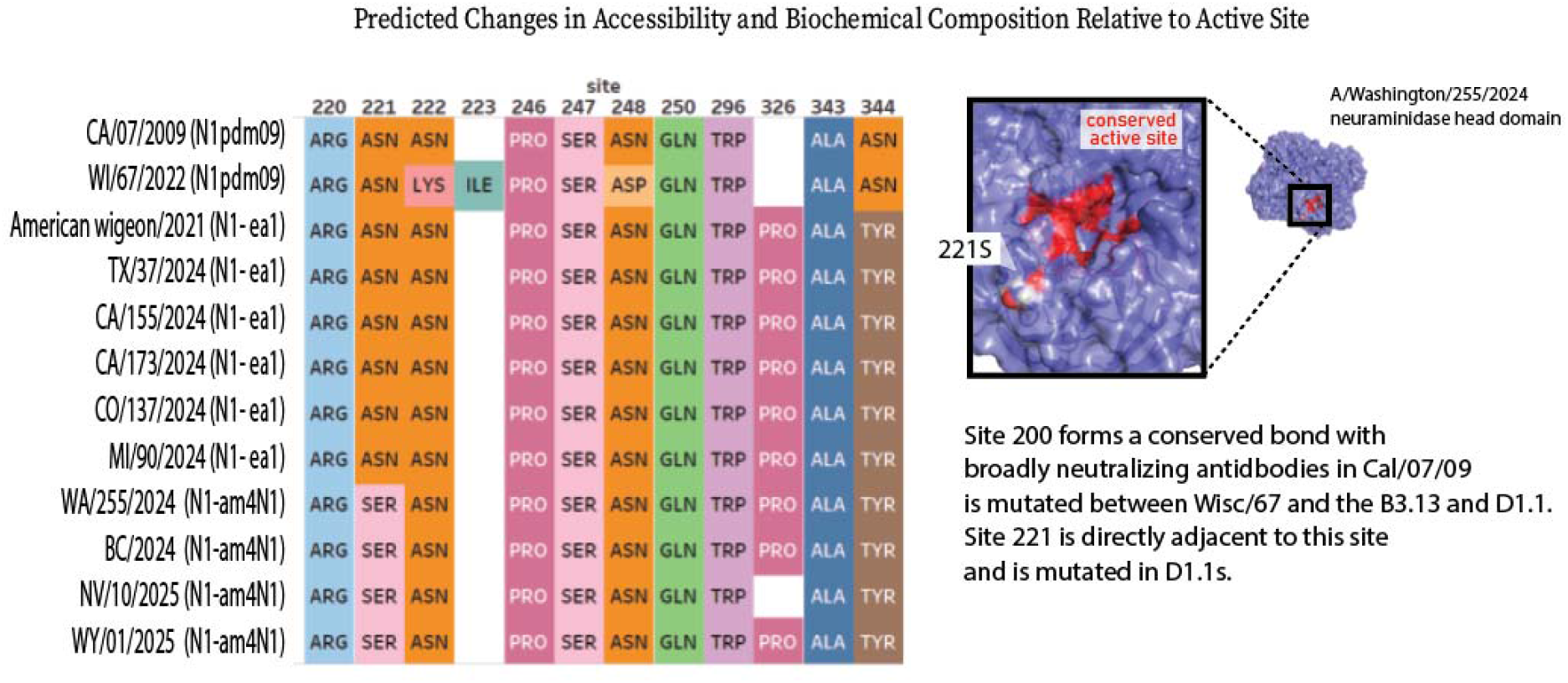
Left) Surface exposed sites (defined as relative surface accessibility of greater than 0.25) within 25 Angstrom (Å) radius of the neuraminidase active site utilizing N1 numbering. Colors indicated differing identities at homologous positions. Right) A/Washington/255/2024 NA structure with the 221S substitution highlighted. Conserved active site residues are colored red.

## Discussion

HPAI A(H5N1) viruses have posed a significant pandemic threat since their emergence in China in the mid-1990s. Within the context of the current North American epizootic, A(H5N1) has shown an ability to diversify rapidly via reassortment. Given this and the high number of documented spillover events recently in North America, understanding the present diversity of reassorted genotypes is critical for the accurate assessment of pandemic risk. The NA genes associated with the newly emergent D1.1 and D1.2 genotypes of clade 2.3.4.4b A(H5N1) in North America are highly divergent from previously observed NAs associated with this HA clade. Before reassorting with clade 2.3.4.4b A(H5) HAs, the am4N1 NA and its progenitors were previously paired with avian A(H1) viruses almost exclusively observed from North America migratory waterfowl. Interestingly, our global survey of N1 diversity showed that this lineage descends from the earliest detections of HPAI from the late 1950s (e.g., A/Chicken/Scotland/1959).

Structurally, NAs appear largely conserved within and across host associations, however, amino acid sequences are divergent between am4N1s and other Eurasian N1s and A(H1N1)pdm09 NAs. Am4N1s have shown seven different instances of NA stalk deletions, with two separate events appearing as convergent for SΔAA:53-78. There is limited evidence to suggest widespread circulation of the deletion variants, as they have been isolated from several wild bird hosts and one wild mammal (Table 1). Typically, stalk deletions in NA are associated with adaptation of HPAI from wild birds to gallinaceous hosts [28]. Indeed, the nodes ancestral to the observed deletions within the tree are usually predicted as being associated with poultry hosts (supplemental data figure 2). As am4N1 and its predecessors have primarily been isolated from wild birds, it is not unexpected that as D1.1 has spread widely among poultry in the US it would pick up deletions, a hallmark of host range expansion [29, 30]. The degree to which these deletions impact viral fitness and functional balance between the HA and NA should be further explored.

Recent studies suggested that previous exposure via vaccination and infection with human-endemic seasonal influenza viruses (e.g., A(H1N1)pdm09) may provide some cross-reactive antibody protection to A(H5N1) viruses [31]. Similarly, La Sage et al. [32] has shown that prior exposure to A(H1N1)pdm09 provide protection from severe disease in ferret models. Previous studies using ferrets inoculated with human seasonal vaccines suggested that this overall protection was primarily driven by immune responses to the N1 NA [33]. To date, no severe disease has been reported in the current North American A(H5N1) outbreak from the genotype B3.13 viruses [9], which carry Eurasian N1s [34]. These N1s are more closely related to contemporary human endemic N1 neuraminidases (i.e., A(H1N1)pdm09) than the am4N1s found in D1.1 viruses (Figure 1). In contrast, viruses of the D1.1 genotype are reported to have caused severe disease requiring hospitalization and two deaths. NA genes have been shown to play a critical role in viral replication (26, 34). Given the highly divergent nature of the NA genes associated with these infections and the potential for reduced antibody cross-recognition of am4N1-NAs, future studies should assess the potential variability of cross-protection afforded by seasonal influenza vaccination. Am4N1 have an amino acid substitution at NA1:N221S, which is near the active site and identified as a site relevant to monoclonal antibody escape [27]. Our surface accessibility mapping shows this site is exposed (Figure 4). Stadlbauer et al. [35] identified that broadly neutralizing antibodies bind to this region and function to block the NA active site especially the area surrounding residue 221 (N2 numbering 220). Stadlbauer’s N1 representative did not include an N1 neuraminidase from the American lineage from which am4N1 descends. Further studies are needed to characterize the degree to which prior N1 infection or vaccination may illicit antibody recognition of am4N1-like NAs generally, and the degree to which amino acid identity may allow for escape.

Pandemic preparedness countermeasures such as vaccines typically rely on comparing host antibody recognition via Hemagglutinin inhibition assays (HAI) as the HA mediates viral attachment and host cell entry and is the primary surface antigen. However, the NA plays a key role in influenza viral replication as well-allowing the release of progeny virions from infected cells [27, 36]. Neuraminidase activity is targeted by numerous pharmaceutical countermeasures including the widely prescribed oseltamivir which inhibits its enzymatic activity [37]. Currently, Neuraminidase inhibiting pharmaceuticals are stockpiled for the event of an influenza pandemic. Signore et al. [8] showed that am4N1 NAs isolated from Canadian chickens harbored an amino acid substitution at NA:H275Y, a known marker for resistance to oseltamivir and showed that the isolate with NA:H275Y had decreased susceptibility to oseltamivir. This clade has not shown onward transmission in the samples analyzed here but continued surveillance for NA:H275Y is needed. Most am4N1 NAs do not carry this amino acid substitution and human isolates tested to date are inhibited well by NAI pharmaceuticals [38].

Another critical aspect of pandemic preparedness is fully understanding the genetic makeup and evolutionary history of pathogen threats. Here we provide a description of the phylogenetics and evolution of am4N1 NAs which have contributed to the ongoing epizootic in wild birds, infected numerous mammals, and caused two human fatalities to date. In addition, we use bioinformatic analysis to predict the structure of diverse NAs and determine how sequence identity changes may have functional implications for am4N1 NAs. Am4N1-like NAs are highly divergent from those previously associated with HPAI A(H5N1) viruses from clade 2.3.4.4b and are phylogenetically distant from NA genes that humans may have immunity to via vaccination and prior infection. Detailed studies describing the human antibody recognition of viruses with the am4N1 protein following seasonal influenza vaccination or infection are needed to fully understand the public health risk posed by D1.1 viruses and its derivate genotypes sharing this NA lineage.

## Acknowledgements

We thank the staff of the Influenza Division and all personnel from the US CDC deployed on the A(H5N1) response. We greatly appreciate the efforts of the staff of the various state and local public health agencies involved in the initial investigation, collection, and detection of positive human samples. This study was enabled by data from GISAID. We thank the USDA, PHA Canada, and all other submitters to GISAID and NCBI for making their data available for study.

## Data Availability

GISAID data used in this study can be found at: EPI_SET_251204qo (https://doi.org/10.55876/gis8.251204qo) for the am4N1 analysis and EPI_SET_260209yr (https://doi.org/10.55876/gis8.260209yr) for the Global N1 analysis. All code is available at: https://github.com/CDCgov/am4N1Phylogenetics. Auspice/Nextstrain JSONs with analytical results are indexed at: https://doi.org/10.7910/DVN/0CMRRX.

## Supplemental Materials

**Supplemental Figure S1.**
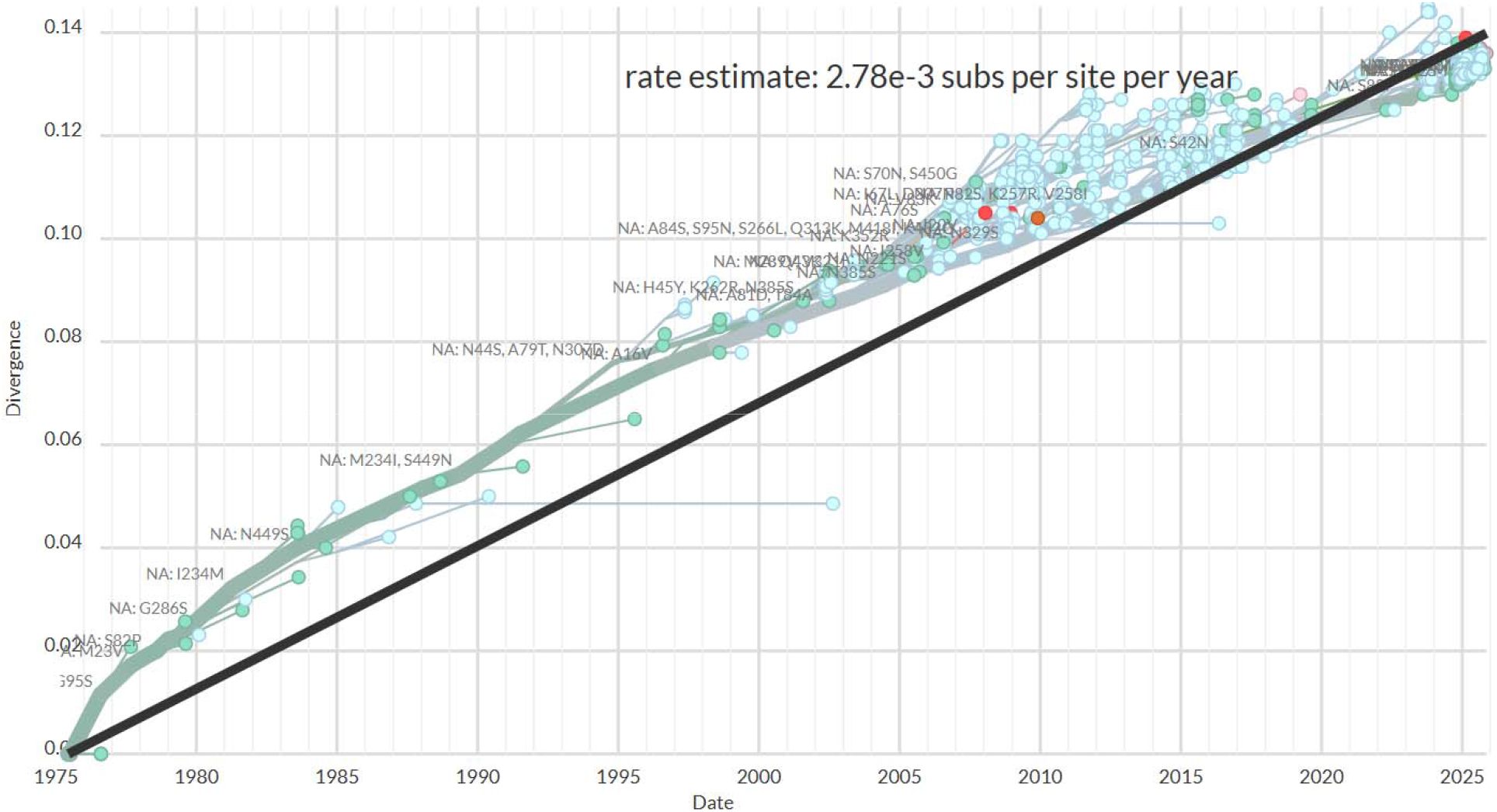
Root to tip regression of am4N1 with major amino acid substitutions shown along tree backbone. Tight correlation suggests the evolution pattern follows a strict molecular clock.

**Supplemental Figure S3.**
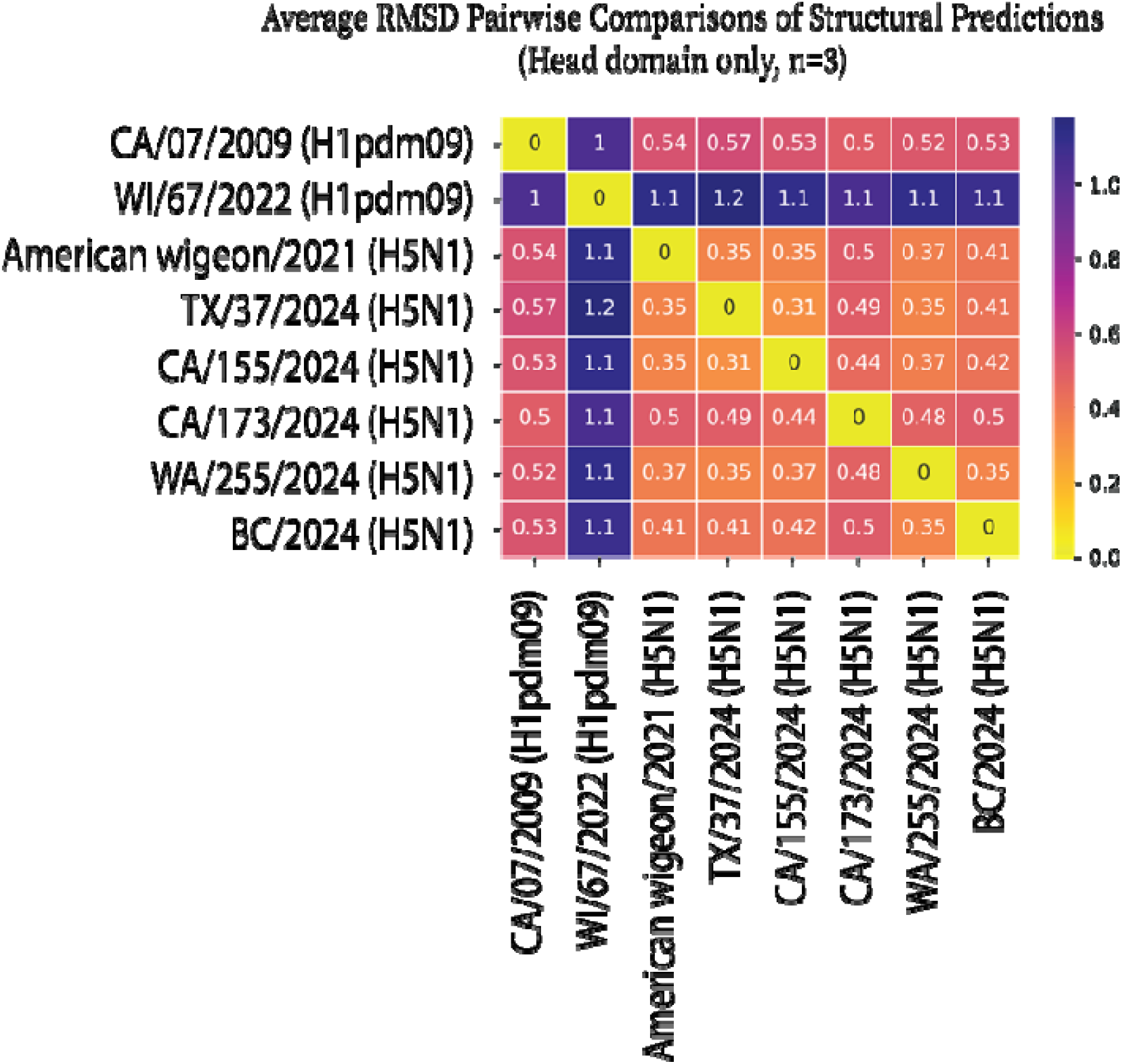
AlphaFold 3 structural predictions were optimized using Rosetta relax and scored using Rosetta scoring algorithms. The RMSD values were averaged across three replicates and plotted in a heat map using the python seaborn library. We excluded sequences with large deletions in the stalk because RMSD are too large to be comparable (>15).

**Supplemental Table S1.**
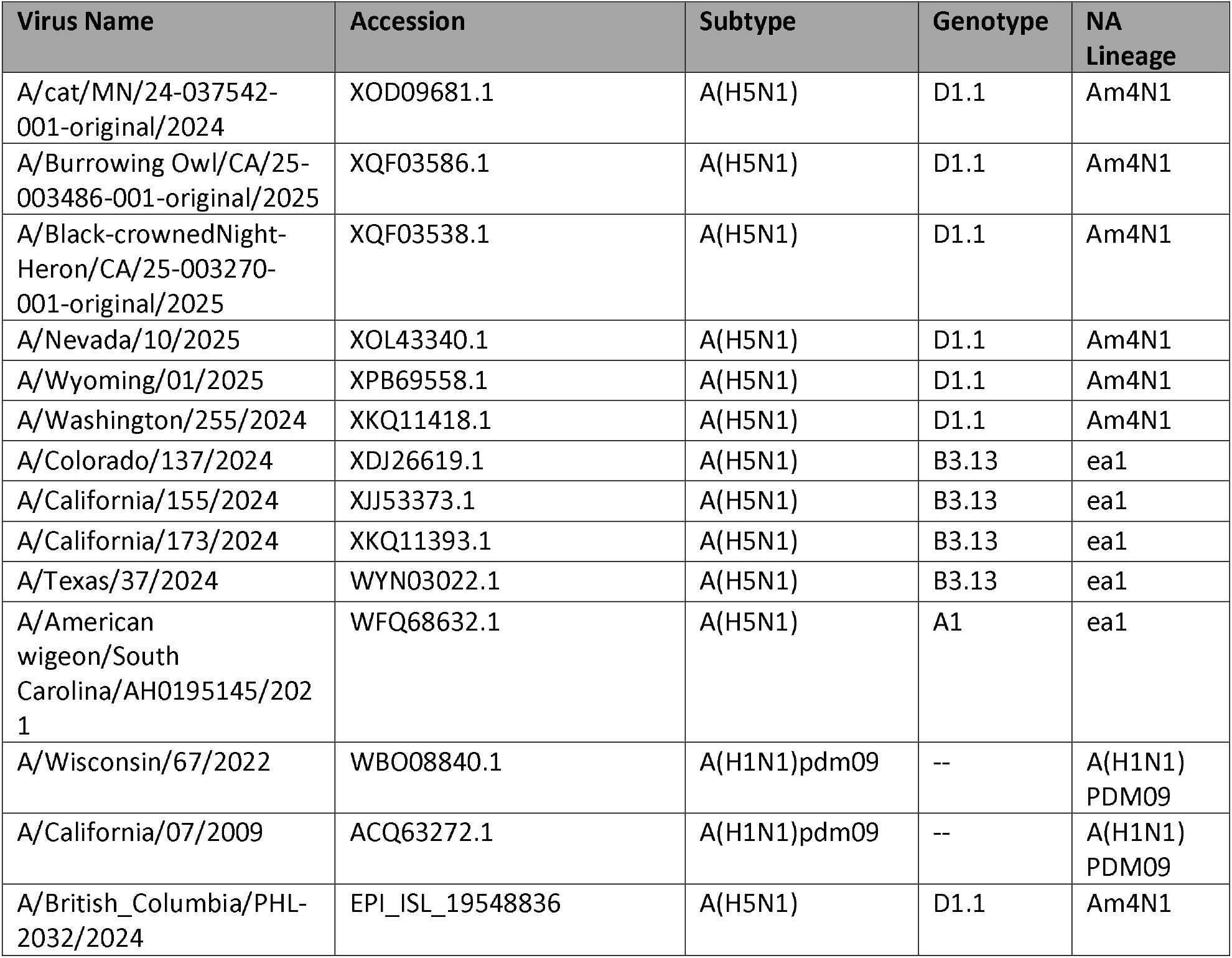
Genbank and GISAID accesssion numbers for amino acid sequences used in the AlphaFold 3 prediction.

| Virus Name | Accession | Subtype | Genotype | NA Lineage |
| --- | --- | --- | --- | --- |
| A/cat/MN/24-037542-001-original/2024 | XOD09681.1 | A(H5N1) | D1.1 | Am4N1 |
| A/Burrowing Owl/CA/25-003486-001-original/2025 | XQF03586.1 | A(H5N1) | D1.1 | Am4N1 |
| A/Black-crownedNight-Heron/CA/25-003270-001-original/2025 | XQF03538.1 | A(H5N1) | D1.1 | Am4N1 |
| A/Nevada/10/2025 | XOL43340.1 | A(H5N1) | D1.1 | Am4N1 |
| A/Wyoming/01/2025 | XPB69558.1 | A(H5N1) | D1.1 | Am4N1 |
| A/Washington/255/2024 | XKQ11418.1 | A(H5N1) | D1.1 | Am4N1 |
| A/Colorado/137/2024 | XDJ26619.1 | A(H5N1) | B3.13 | ea1 |
| A/California/155/2024 | XJJ53373.1 | A(H5N1) | B3.13 | ea1 |
| A/California/173/2024 | XKQ11393.1 | A(H5N1) | B3.13 | ea1 |
| A/Texas/37/2024 | WYN03022.1 | A(H5N1) | B3.13 | ea1 |
| A/American wigeon/South Carolina/AH0195145/2021 | WFQ68632.1 | A(H5N1) | A1 | ea1 |
| A/Wisconsin/67/2022 | WBO08840.1 | A(H1N1)pdm09 | -- | A(H1N1) PDM09 |
| A/California/07/2009 | ACQ63272.1 | A(H1N1)pdm09 | -- | A(H1N1) PDM09 |
| A/British_Columbia/PHL-2032/2024 | EPI_ISL_19548836 | A(H5N1) | D1.1 | Am4N1 |

